# Latent EBV impairs immune cell signaling and enhances the efficacy of anti-CD3 mAb in Type 1 Diabetes

**DOI:** 10.1101/2023.07.11.23292344

**Authors:** Ana Lledo Delgado, Paula Preston-Hurlburt, Noha Lim, Tomokazu S. Sumida, S. Alice Long, James McNamara, Elisavet Serti, Lauren Higdon, Kevan C. Herold

**Affiliations:** Department of Immunobiology, Yale University School of Medicine, New Haven, United States of America; Immune Tolerance Network, Bethesda, MD 20814, USA; Center for Translational Immunology, Benaroya Research Institute at Virginia Mason, Seattle, WA, USA; Autoimmunity and Mucosal Immunology Branch, Division of Allergy, Immunology and Transplantation, National Institute of Allergy and Infectious Diseases, National Institutes of Health, Bethesda, MD, USA

## Abstract

Teplizumab has been approved for the delay of the onset of type 1 diabetes and may modulate new onset disease. We found that patients who were EBV positive at baseline had a more robust response to drug in two clinical trials and therefore postulated that latent virus has general effects in modifying immune responses. We compared the phenotypes, transcriptomes, and development of peripheral blood cells before and after teplizumab treatment. Higher number of Tregs and partially exhausted CD8^+^ T cells were found in EBV seropositive individuals at the baseline in the TN10 trial and AbATE trial. Single cell transcriptomics and functional assays identified downregulation of the T cell receptor and other signaling pathways before treatment. Impairments in function of adaptive immune cells were enhanced by teplizumab treatment in EBV seropositive individuals. Our data indicate that EBV can impair signaling pathways generally in immune cells, that broadly redirect cell differentiation.

## Introduction

Teplizumab, the FcR non-binding humanized anti-CD3 mAb was the first biologic approved for delay of autoimmune disease (type 1 diabetes) and it may modulate new onset disease. However, the clinical responses to teplizumab are varied and there is little known about the determinants of these responses. Latent viruses, such as Epstein Barr virus (EBV) and Cytomegalovirus (CMV) may modify human immune cells and are postulated to play a role in autoimmunity. It has been suggested that they have direct causative roles in some autoimmune diseases such as multiple sclerosis (MS), rheumatoid arthritis (RA), systemic lupus erythematosus (SLE), Sjogren’s syndrome, and potentially others^1–5^,but the ways in which it affects these diseases are not understood. Nearly all patients with MS have previous exposure to EBV and EBV seroconversion has been shown to associate with the onset of disease. Cross reactivity between antibodies that bind Epstein–Barr nuclear antigen 1 (EBNA1) and the central nervous system protein glial cell adhesion molecule has been discovered in patients with MS^6^.

In addition to direct recognition of viral antigens, latent viruses may affect human responses in other ways. Among genetically identical twins, exposure to CMV was shown to modify the T cell repertoire^7^. Viruses establish latency in and modify affected tissues. EBV and CMV both establish latency in immune cells and therefore can change them directly. Identifying the effects of EBV and CMV on human autoimmune responses has been challenging since cause and effect cannot be clearly established. Many patients have already been exposed to these viruses by the time they present with autoimmunity-most of the patients are adults where the seroprevalence to the viruses is very high. Model systems in which direct relationships between latent viruses and immune responses can be identified are limited since human EBV does not infect mice, and mice that can be infected with counterparts do not develop autoimmune diseases spontaneously.

Previous investigations of the mechanisms of action of teplizumab have shown that the drug caused partial-exhaustion signature in CD8^+^ T cells among drug-treated participants characterized by an increased frequency of CD8^+^ cells expressing the transcription factor EOMES, and KLRG1 and TIGIT ^8–11^. Comparisons of responses to biologics, in patients with and without these latent viruses, can give insights into the immune effects of the viruses and the biologics in ways that are not possible with observational studies. Combined data from clinical trials of anti-CD3 mAb gave us the opportunity to understand the role of latent viruses on human immune responses, since among the participants in the studies, the majority of whom were children, only about half had previously been exposed to EBV and a smaller proportion to CMV. Therefore, to understand how these viruses affect human immune responses we compared them in patients who had or had not previously been infected with EBV who were treated with teplizumab in two independent clinical studies. We identified changes in CD4^+^, Tregs, and CD8^+^ T cells in EBV seropositive vs seronegative individuals at the baseline and found more robust clinical responses to teplizumab in the seropositive study participants. Antigen receptor signaling pathways were reduced in T and B cells in EBV seropositive individuals, but the induction of T cell exhaustion was enhanced by teplizumab. Collectively, these findings before and after treatment with teplizumab suggest a pervasive effect of EBV on adaptive immune cells. The significance of latent EBV on clinical outcomes, such as autoimmunity and tumorigenesis, may vary by the cells engaged in the disease specific responses.

## Results

### Differences in multiple immune cell subsets associated with EBV serostatus

To identify the effects of prior exposure to EBV or CMV on immune cells, we compared the phenotypes of cells from the peripheral blood (PBMCs) of the 76 participants who were at high risk for development of clinical T1D in the TN10 trial, and 75 participants recently diagnosed with T1D in the AbATE trial, two clinical studies of teplizumab. The TN10 trial evaluated the time from treatment, with a single 14 day course of teplizumab vs placebo, until the diagnosis with clinical Stage 3 T1D. Of the 76 participants, 18 were EBV seropositive (Table 1). The frequency of positive tests for EBV was closely associated with age quartile (Chi-squared p<0.0001). The AbATE trial evaluated the effects of two 14 courses of teplizumab on stimulated C-peptide responses at 2 years. At enrollment, 35% were EBV seropositive (8/23 in the control and 18/52 in teplizumab arms). The PBMCs were analyzed with 260 parameters evaluating the frequency and intensity of expression of markers on T, Tregs, NK and NKT cells (Supplemental Table 1, gating in ^8, 12^). We found significant differences in the expression of 76 markers (p<0.05) on cell subsets when the phenotypes of cells from EBV seropositive and EBV seronegative patients were compared, 11 after FDR correction. (Figure 1A). The differences between the EBV seropositive and seronegative participants were in markers expressed generally on CD8^+^, CD4^+^, and Tregs.

**Figure 1.**
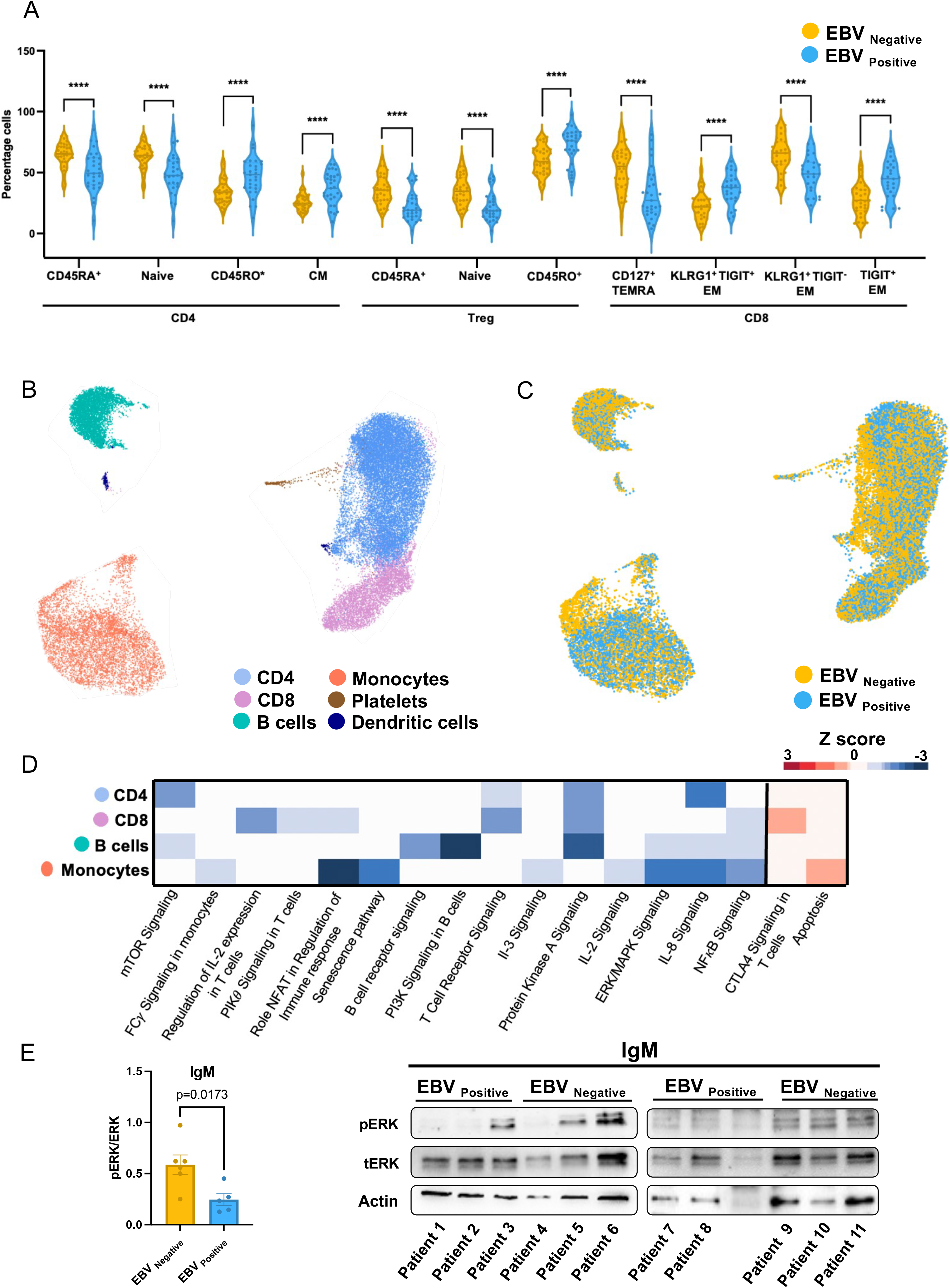
Changes in immune cell subsets at the baseline among EBV positive or negative individuals. (A) Violin plots showing the percentages of populations from the TN10 trial analyzed by flow cytometry and found to be significantly different after FDR correction. Colors identify EBV serological status (n=29 EBV seropositive, n=34 EBV seronegative). (B) UMAP visualization of the clusters at the baseline. Points represent individual cells and color denote cluster classification as labeled (n=6 EBV seropositive and n=8 EBV seronegative). (C) UMAP visualization of the cells at the baseline. Points represent individual cells and color denote EBV serological status. (D) Heatmap showing the Z score of the significant pathways (p<0.05 after FDR correction) in the major clusters at the baseline. Pathways were inferred based on the DEGs between EBV positive and EBV negative in each cell subset using IPA software. Blue and red scale denote grades of prediction of downregulation or upregulation of the pathways based on the Z score. (E) Western blot showing phosphorylation of ERK in B cells after stimulation with anti-human IgM. The ratio of pERK/totalERK was corrected for loading (actin). The levels of pERK are decreased in the EBV seropositive (n=6) vs EBV seronegative (n=5) individuals (*p=0.01). CM:central memory; TEMRA: T effector memory RA; EM: effector memory

The co-infection with CMV, another common latent virus may have affected the phenotypes of the cells. There were 36 parameters that were significantly different (p<0.05) when we compared the participants who were EBV seropositive only (n=17) to those who were EBV and CMV seropositive (n=11) (Supplemental Table 2)(but none of these differences met statistical significance after correction for multiple comparisons (two-stage step-up method).

To further understand the differences among immune cell subsets in the EBV seropositive and seronegative individuals, we compared transcriptional profiles of PBMCs by single-cell RNA-seq (scRNA-seq) with 10X Chromium 3′ sequencing platform. We annotated five major clusters based on the expression of key identity genes: CD8^+^ T cells (*CD8A*^hi^*, CD3D*^hi^*, CD3E*^hi^), CD4^+^ T cells (*CD8A*^-^, *CD3D*^hi^, *CD3E*^hi^), B cells (*CD79A*^hi^, *MS4A1*^hi^), monocytes (*CD14*^hi^, *FCGR3A*^hi^) dendritic (*PLD4*^hi^, *LILRA4*^hi^) and platelets (*CD3*^-^ *PPBP*^+^) (Figure 1B and Supplemental Figure 1). We found significant differences in the major cell subsets based on the EBV serological status. We identified 57, 41, 157, 35, 123 and 2 genes that were differentially expressed in the CD4^+^, CD8^+^, Treg, B cells, monocytes, and dendritic cells at the baseline (Figure 1C and Supplemental Table 3). Figure 1D shows pathways that were differentially expressed using IPA analysis (Figure 1D and Supplemental Table 4). Consistent with our findings by flow cytometry, in CD8^+^

T cells, there was increased expression of pathways related to exhaustion (increased CTLA4 signaling in CD8^+^ T cells (p=2.96E-05)) but, more generally, reduced expression of genes in antigen receptor signaling among CD4^+^, CD8^+^ T, and B cells. (e.g. PI3K signaling in B cells (p=3.72E-06), protein kinase A signaling in CD4^+^, CD8^+^ and B cells (p=1.56E-04, p=1.37E-04, p=2.43E-04), NFκB signaling in CD8^+^ T cells and B cells (p=3.23E-04 and p=1.45E-02) and reduced mTOR signaling in CD4^+^ T cells (p=2.48E-02) and others.

Among B cells, we found significant differences in B cell receptor and ERK/MAPK signaling (Figure 1E). To validate this finding, we compared ERK phosphorylation in B cells from a separate group of EBV seropositive or seronegative individuals with T1D. Following IgM cross-linking, there was higher levels of ERK phosphorylation in 6 EBV seronegative vs 5 EBV seropositive donors (p=0.01).

### EBV seropositive patients have improved clinical responses to teplizumab treatment

These findings *ex vivo* distinguished phenotypic and transcriptional differences in cell subsets among EBV seropositive and seronegative individuals that may affect their functional responses *in vivo*. To understand the significance of these differences, we compared the clinical responses to teplizumab treatment in the two clinical studies based on EBV serostatus at the time of drug treatment (Table 1).

Among those who were EBV seropositive the median time to diagnosis with Stage 3 T1D was delayed significantly with teplizumab treatment (median times from study entry until the time to diagnosis of Stage 3 diabetes: 35.5 months (placebo, n=16) vs 86.9 months (teplizumab, n=18), p=0.038 Logrank test after Sidak’s correction) (Figure 2A, B). In the EBV seronegative participants there was also a delay in the time to diagnosis with Stage 3 T1D, but the difference was not statistically significant (12.0 months (placebo, n=16) vs 38.0 months (teplizumab, n=26), p=0.719). At the end of the observation period, 12.5% (2 out of 16) of the placebo treated EBV seronegative and seropositive individuals were diabetes free whereas 26.9% EBV seronegative (7 out of 26 Fisher’s exact test, p=0.44 vs placebo) and 50% (9 out of 18) were EBV seropositive individuals (p=0.03) treated with teplizumab were diabetes free. Among patients who were EBV seropositive at study entry (n=34), 8 of those in the teplizumab group had detectable viral loads at weeks 3-6, that cleared spontaneously. However, the responses to teplizumab were similar in those with and without detectable EBV viral loads.

**Figure 2.**
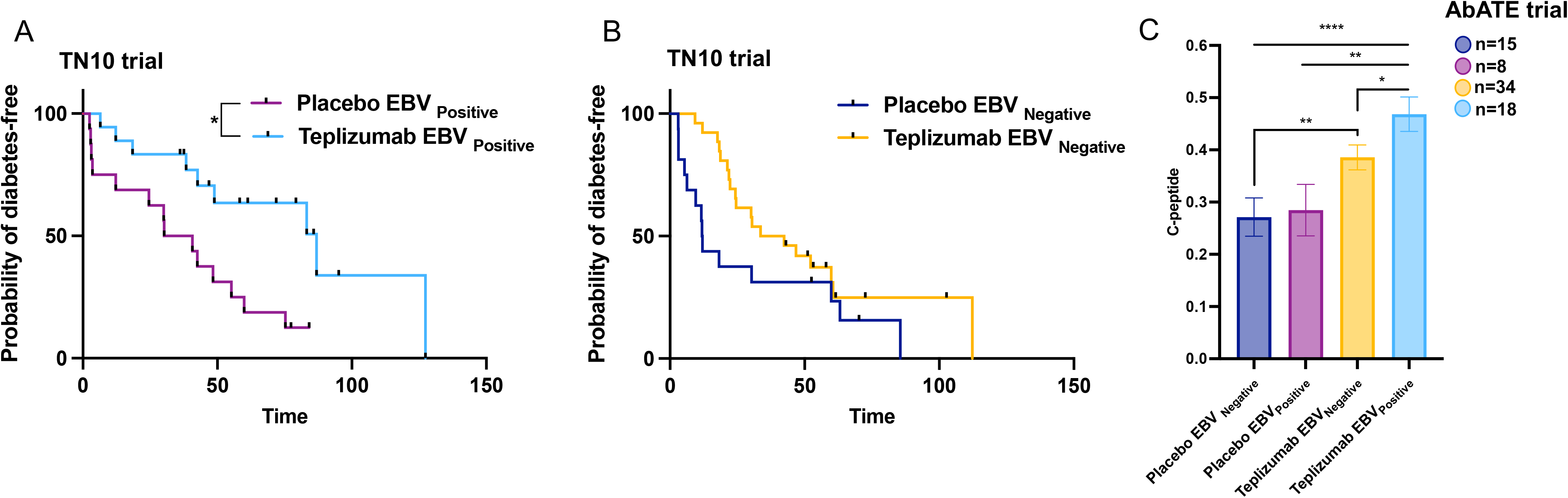
Effects of EBV serostatus on clinical responses to teplizumab. (A-B) Kaplan Meier curve showing the progression from Stage 2 T1D to Stage 3 T1D in TN10 study participants who were (A) EBV seropositive at enrollment (n=18, teplizumab, n=16 placebo, p=0.038 Logrank test after Sidak’s correction) or (B) EBV seronegative at enrollment (n=26 teplizumab, 16 placebo, p=0.719). (C) Effects of EBV serostatus in C-peptide responses in the AbATE trial (n=15 placebo EBV seronegative, n=8 placebo EBV seropositive, n=34 treatment EBV seronegative, n=18 treatment EBV seropositive). The data are from a mixed linear model with correction for the baseline response, and shows the mean±95% CI for each treatment/serostatus group. (*p<0.05, **p<0.01, ****p<0.0001)

The number of participants who were CMV seropositive and either EBV seronegative or EBV seropositive (n=5 and 12) limited our ability to assess the effects of CMV serostatus on the response to teplizumab. However, the same trend of increased median time to progression to Stage 3 T1D was found in the dual positive participants when placebo treated (median = 55 months) and teplizumab treated (median 83 months) were compared.

We confirmed these clinical findings in a second trial that evaluated the effects of teplizumab on stimulated C-peptide responses over 2 years, in patients who had new onset clinical (Stage 3) T1D, (ITN027AI, “AbATE”) (Table 1). Teplizumab treatment improved the stimulated C-peptide levels (vs control) in both the EBV seronegative (difference of least square means (LSM)=0.113±0.043, p=0.0086) and EBV seropositive participants (difference LSMs = 0.185±0.058, p=0.0018) but the C-peptide was significantly greater in the EBV seropositive vs EBV seronegative teplizumab treated individuals (difference LSMs = 0.091±0.04, p=0.024) (Figure 2C).

### EBV seropositive patients have increased expression of exhaustion markers on CD8^+^ T cells with teplizumab treatment

Previously, teplizumab was shown to induce “partial exhaustion” of CD8^+^ T cells which was identified by an increase in the expression of EOMES^+^ and frequency of KLRG1^+^TIGIT^+^ CD8^+^ cells^11^. We compared, by flow cytometry, EOMES expression and the frequency of these cells over time after teplizumab treatment in the two trials. In the TN10 study, there was a higher frequency of the EOMES^+^ CD8^+^ T effector memory RA (TEMRA) cells before study drug treatment (p=0.0004) in EBV seropositive individuals. Moreover, the frequency of EOMES^+^ CD8^+^ T cells, was increased in CD8^+^ central memory (CM) (LS means: 44.2+2.35% vs 35.5+2.07%, p=0.007) (Figure 3A), and effector memory RA cells (EMRA) (LS means: 75.1+2.4% vs 54.8+2.11%, p<0.0001), in the EBV seropositive vs seronegative participants after treatment. There was a corresponding increased gene expression of EOMES in CD8^+^ T cells (Figure 3B, C) in scRNAseq (n=4 EBV seropositive, n=4 EBV seronegative). In addition, KLRG1^+^TIGIT^+^ expression was greater on CD8^+^ T cells at the baseline (Figure 3E, p=0.012) and was also increased after treatment on CD8^+^ CM (LS means 35.8+2.91% vs 18.8+2.51%, p<0.0001), effector memory (EM) (LS means: 49.7+3.3% vs 22.7+2.85%, p<0.0001), and EMRA cells (LS means: 58.9+3.78 vs 37.9+3.26, p<0.001). The overall increased of KLRG1^+^ TIGIT^+^ was confirmed by scRNAseq in the TN10 trial at 3 months (Figure 3F). In the AbATE trial, the same observations were confirmed, the frequency of CD8^+^ EOMES^+^ (p=0.005) and KLRG1^+^ TIGIT^+^ (p=0.036) T cells were higher in the EBV seropositive individuals (Figure 3D, G).

**Figure 3.**
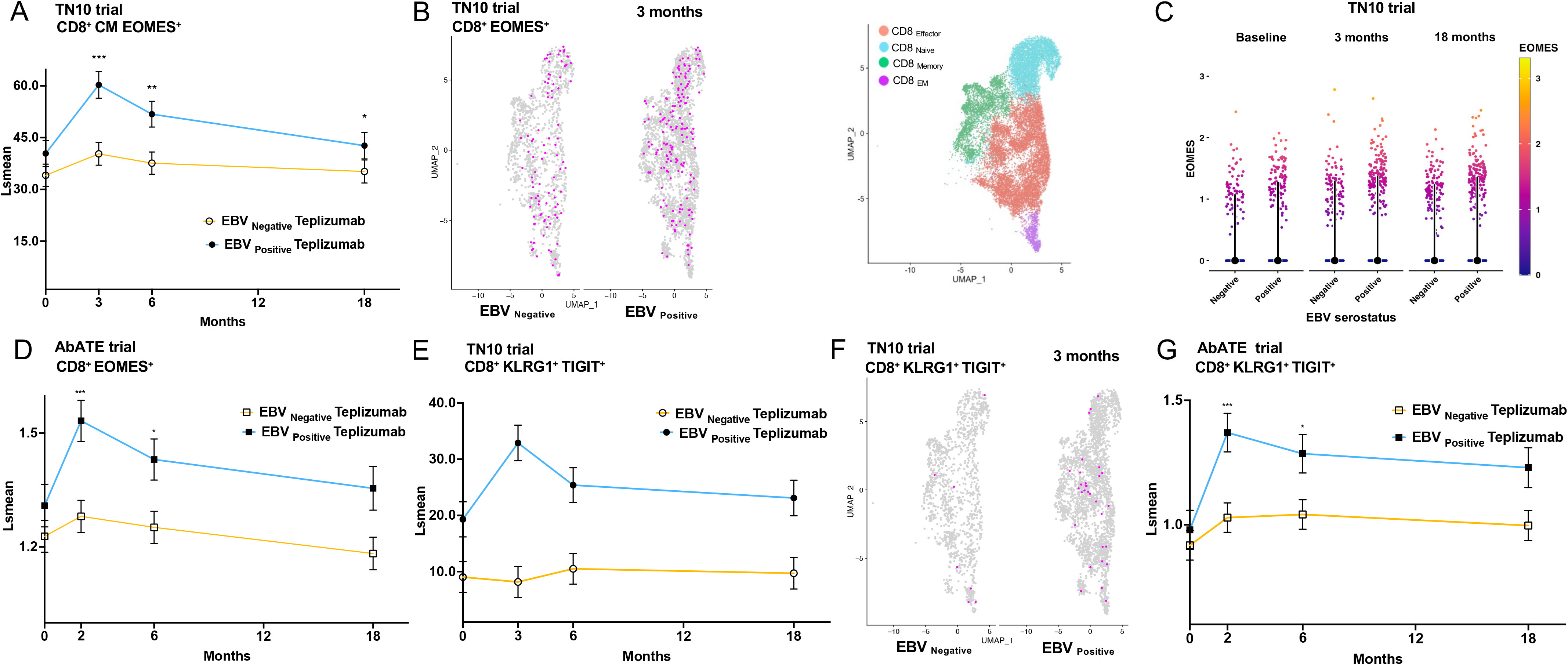
Induction of “partially exhausted” CD8 T cells with teplizumab in clinical trial participants. (A-C) Increased frequency of CD8^+^EOMES^+^ T cells among CD8^+^ CM T cells in the EBV seropositive (n=16) vs EBV seronegative (n=22) teplizumab treated in the TN10 trial by flow cytometry, (A, p=0.0068) and among CD8^+^ T cells by single cell RNAseq (B, C, color denotes levels of gene expression and points represent individual cells, n=4 in each EBV group). (D) Increased frequency of CD8^+^EOMES^+^ T cells in the EBV seropositive (n=10) vs EBV seronegative (n=18) teplizumab treated in the AbATE trial by flow cytometry, (p=0.005). (E-F) Increased frequency of KLRG1^+^ TIGIT^+^ CD8^+^ T cells in the TN10 trial among EBV seropositive (n=16) vs EBV seronegative (n=22) participants by flow cytometry at the baseline (p=0.029) and with teplizumab treatment (E, p<0.0001) and by gene expression (F, points represent individual cells, n=4 in each EBV group). (G) Increased frequency of KLRG1^+^TIGIT^+^ CD8^+^ T cells (p=0.036) in the AbATE trial. CM: central memory

### Interactions between EBV serostatus and teplizumab responses in patients

These findings, from two independent clinical trials of teplizumab, showed increased frequencies of CD8^+^ T cells that are purported to mediate the biologic effects of the drug in the EBV seropositive participants. However, our flow cytometry analysis indicated there were effects of EBV on other cells prior to teplizumab treatment. We therefore compared the changes in the frequency and transcriptomes of immune cells of 4 patients EBV seropositive vs 4 patients EBV seronegative over time after teplizumab treatment in the EBV seropositive and EBV seronegative participants in the TN10 trial to identify how the effects on CD8^+^ and other cells may contribute to the improved efficacy. The samples were obtained approximately 3 months (first visit) and 18-24 months (last visit) after study drug).

We did not find a significant difference in the frequencies of CD8^+^, CD4^+^ T cells, B cells, monocytes, or dendritic cells overall (Supplemental Figure 2).. We further analyzed the cell subsets by scRNA-seq using transcriptomic markers to identify them described in Supplemental Figure 1. The UMAPs showing the subclustering of the cells among EBV seropostive and seronegative patients are shown in Supplemental Figure 3 A-H. There were trends of differences in the frequency of cell subsets but the differences in the number of cells did not reach statistical significance (Supplemental Figure 3I-L).

We performed pathway analyses (IPA) and compared gene expression between the EBV seropositive and seronegative patients who were treated with teplizumab at different timepoints in the cell subsets defined by expression of these same genes (Figure 4, Supplemental Table 5). There was reduced expression of genes in the NFκB and T cell receptor signaling pathways (p=3.99E-04 and p=5.11E-09) respectively, in CD8^+^ T effector cells at the baseline in the EBV seropositive patients, but at 3 and 18 months after teplizumab treatment, these and other pathways showed further reduction in expression in the EBV seropositive patients. In addition, in CD8^+^ effector cells, pathways of cytokine signaling (STAT3 (p=1.96E-03), IL-2 signaling (p=9.28E-09)) as well as ERK/MAPK (p=9.39E-05) and T cell receptor signaling (p=1.1E-14) showed reduction at the 18 month visit (Figure 4A, B, C). (p values in Supplemental Table 6). Consistent with these findings by scRNA-seq, of reduced T cell signaling in the EBV seropositive individuals, we found that the MFI of CD3 was reduced on CD8^+^ CM T cells in the EBV seropositive vs seronegative patients at the baseline (p=0.007) and after teplizumab treatment (LS means: 13047+278 vs 14299+245, p=0.0009) (Figure 4D).

**Figure 4.**
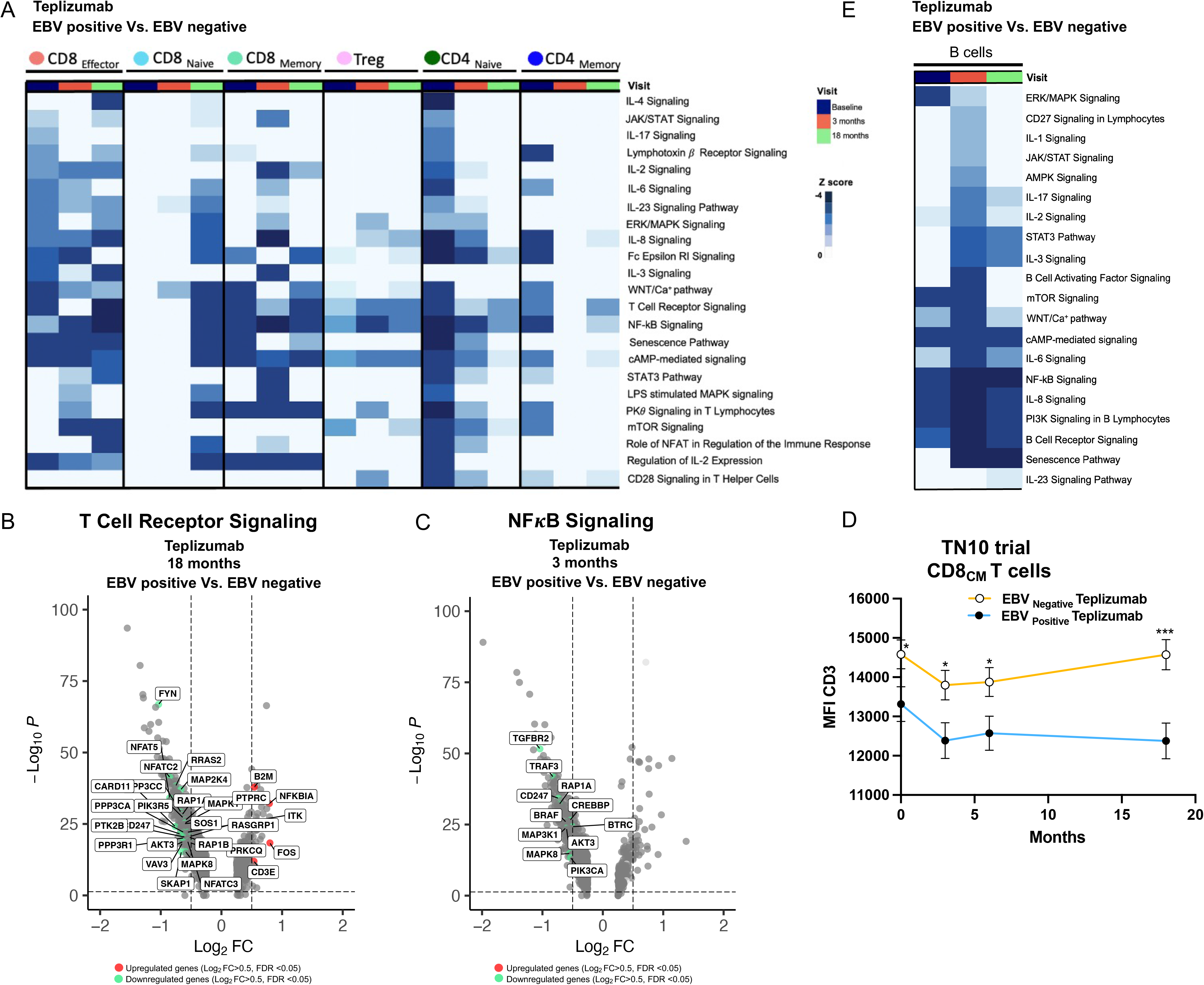
Transcriptional changes and pathway analysis in teplizumab treated patients in the TN10 trial who are EBV seropositive vs EBV seronegative. (A) Heatmap showing the Z score of the pathways with significant differences in the CD8^+^ and CD4^+^ T cells clusters at the baseline, first visit (three months) and last visit (eighteen months) (n=4 EBV seropositive, n=4 EBV seronegative). The pathways were inferred based on the DEGs between EBV positive and EBV negative in the teplizumab group in each timepoint. Blue scale denotes grades of prediction of downregulation of the pathways based on the Z score. (B) Volcano plot visualization of the differentially expressed genes (DEGs) in EBV positive vs negative related to the T cell receptor pathway in the CD8 effector cluster at the last visit (eighteen months), and (C) differentially expressed genes (DEGs) in EBV positive vs negative related to the NF*k*B in the Treg cluster at the first visit (three months). (D) Flow cytometry data from the TN10 trial showing the differences in the CD3 expression in the CD8^+^ CM between EBV seropositive (n=16) and seronegative (n=22) individuals. (E) Heatmap showing the Z score of the significant pathways (p<0.05) in the B cells cluster at the baseline, first visit (3 months) and last visit (18 months). CM: central memory.

T cell receptor signaling (p=1.91E-11) mTOR signaling (p=8.03E-06) and other pathways in CD4^+^ Naive T cells showed the greatest reduction at the baseline. Among B cells, we found significant differences at the first and last visit in cytokine signaling pathways and in B cell receptor and ERK/MAPK signaling (Figure 4E). All these data together suggest a downregulation of the pathways related to cytokine and activation signaling in EBV positive in comparison with EBV negative maintained under teplizumab treatment.

### EBV serostatus modifies T cell differentiation

To understand how T cell development differed between EBV seropositive and EBV seronegative patients in the TN10 trial, we performed a pseudotime analysis comparing the trajectories of T cells from EBV seropositive vs seronegative TN10 study participants treated with teplizumab. UMAPs were prepared to identify different states of cell differentiation that differed between the EBV seropositive and seronegative patients (Figure 5). We compared this analysis at 18 months after drug to understand how EBV serostatus may change the trajectory of T cells after teplizumab and may lead to long term responses.

**Figure 5.**
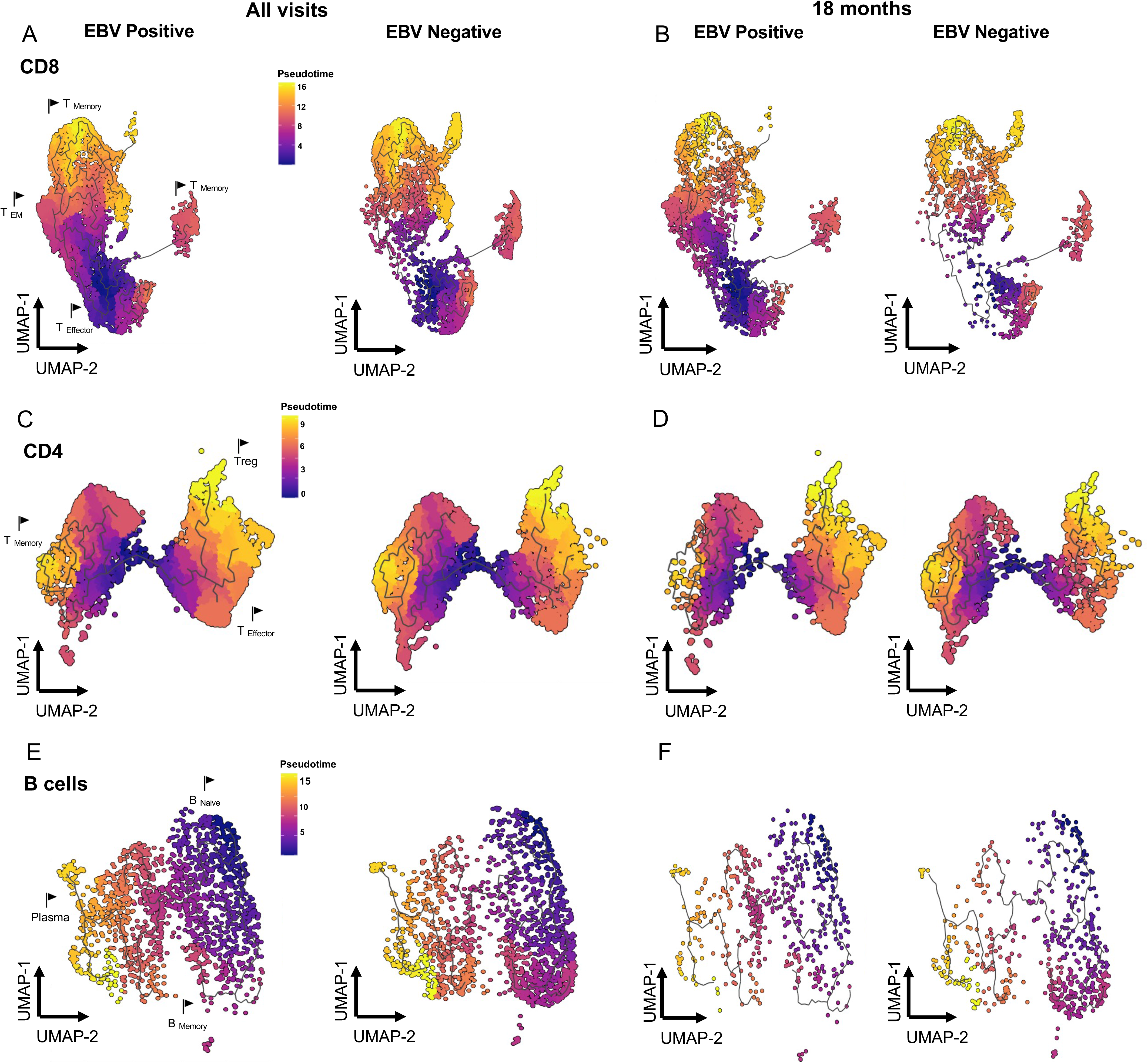
Pseudotime analysis of CD8^+^, CD4^+^ and B cells in the TN10 trial. (A-B) UMAPs showing the pseudotime analysis of CD8^+^ T cells in the EBV seropositive and EBV seronegative in teplizumab treated participants (A) at all of the visits and (B) at 18 months (n=4 EBV seropositive n=4 EBV seronegative). (C-D) UMAPs showing the pseudotime analysis CD4^+^ T cells in the EBV seropositive vs EBV seronegative participants (C) at all of the visits and (D) at 18 months. (E-F) UMAPs showing the different stages of differentiation of B cells in the EBV seropositive and EBV seronegative participants (E) at all of the visits and (F) at 18 months. Transcript dynamics are illustrated by the color of the pseudotime. The location of transcriptional signatures for the major cell states identified are indicated by markers on pseudotime visualizations (T _Memory,_ T_EM_, T_Effector_, Treg, B _Naive_, Plasmablast, B _Memory_).

There were differences in the trajectories of CD8^+^, CD4^+^ T and B cells between the EBV seropositive and seronegative participants at 18 months (Figure 5, and Supplemental Figure 4, 5, 6). In CD8^+^ T cells, we found expression of genes related to exhaustion pathways and inhibition of the immune system in EBV seropositive individuals at earlier stages of the pseudotime. *IL10RA*^+^ (p=3.59E-05), *TIGIT*^+^ (p= 6.35E-28), *LAG3*^+^(p= 0.00) were enriched in the EBV seropositive teplizumab treated patients and maintained at 18 months (Figure 5A-B and Supplemental Figure 4).

In CD4^+^ T cells, *LEF1* was among the top differentially expressed gene in CD4^+^ memory cells (p=2.82E-06 at 3 months) (Supplemental Table 5) between EBV positive and negative. We found differences in the expression across the pseudotime in LEF1, but also in the expression of *TCF7* (p<0.001), *CCR7* (p<0 p<0.001), *IL7R* (p<0.001). Other genes related to the differentiation of Tregs (*CTLA4,* p<0.001) or related to cytotoxic CD4^+^ T cells (*NKG7, GZMK,* p<0.001 *respectively*) were found in early stages of differentiation in EBV positive individuals in comparison to EBV negative at the 18 months (Figure 5 C, D and Supplemental Figure 5). B cells in EBV negative patients showed increased differentiation into B memory and plasmablasts (BANK1^+^, MZB1^+^, CXCR5^+^) at 18 months after teplizumab (Figure 5E-F and Supplemental Figure 6).

## Discussion

By studying the role of EBV in patients treated with teplizumab, we have identified novel immune modulatory effects of the virus and its synergistic interaction with the anti-CD3 mAb teplizumab. EBV has been incriminated as a cause of several autoimmune diseases, but it has largely been considered a target rather than a modifier of pathologic immune responses. We hypothesized that EBV may have effects beyond just the relatively small population of antigen specific CD8^+^ T cells and therefore undertook a comparison of immune cells in individuals who are EBV seropositive and EBV seronegative and compared their responses to teplizumab treatment. Compared to EBV seronegative patients, those with prior EBV infection showed changes in CD4^+^, CD8^+^, Tregs and B cells in PBMCs. The expression of genes that define transcriptional pathways were reduced generally which was confirmed by analyzing phosphorylation of ERK in B cells that were triggered by the BCR. Since the significance of immune modifiers may not be clear under steady state conditions, we took advantage of the disturbances with an immune therapy to identify the effects of latent EBV. In two independently conducted clinical studies of teplizumab, that enrolled patients with new onset or risk for T1D, there was a more robust clinical effect of the drug in the EBV seropositive vs seronegative study participants. In addition, in the EBV seropositive patients, there was an increased frequency of cells with phenotypic and transcriptional functional features of exhaustion, that had previously been found in teplizumab treated patients and had been associated with clinical responses^13^. When we compared the effects of EBV serostatus on the responses to teplizumab over time, we found that there was further inhibition of cell activation pathways which included reduced signaling of the T cell receptors on CD4^+^ and CD8^+^ T cells as well as B cells particularly at the 18 months visit after drug treatment – when the numbers of circulating immune cells were normal. Thus, after teplizumab treatment, there was synergy between functional consequences of latent EBV and the inhibitory activities of the mAb, and these effects persisted. A trajectory analysis identified different cell differentiation states at the 18 months visit that involved transcription of genes associated with cell cytotoxicity and exhaustion. We conclude that latent EBV has broad effects on immune cells and change the differentiation of immune cells in response to disturbances. As a result, latent EBV may affect immune responses that may underlie pathologic or protective immunity.

In a prior study, with another anti-CD3 mAb (otelixizumab), treatment was shown to cause reactivation of latent EBV and there was an increase in the frequency of EBV-reactive CD8^+^ T cells^14^. However, in the TN10 study, the effects of EBV did not appear to require clinical EBV reactivation as detected by EBV viral loads in the peripheral blood, nor were they specific for EBV reactive CD8^+^ T cells since multiple cell types were affected. Of the 18 EBV seropositive participants in the TN10 trial, only 8 had detectable viral loads that were detected at the sampling time point (at 8 weeks), but we did not identify significant differences in clinical outcomes between those who did and did not have detectable viral loads. It is possible that there were increased viral loads that were missed in some patients or at locations such as the nasopharynx or gut where EBV is latent, because of our limited sampling. Furthermore, the viral loads may have returned to normal levels by the time they were measured at 8 weeks after enrollment. It is unlikely that activation of the EBV-specific CD8^+^ T cells or even the EBV-reactive CD4^+^ T cells can account for the improved clinical responses that we found in the two trials or the long-term effects after viral loads had been cleared and viral loads had returned to normal. Importantly, our findings do not reflect a non-specific effect of latent DNA viruses on human immune responses since the effects of teplizumab were either absent or the opposite in individuals who were CMV seropositive. This implies there are specific mechanisms associated with EBV latency and more general effects on immune cells that can modify immune responses.

In addition, the effects of EBV are not the result of direct viral infection of cells. Among resting memory B cells, the site of EBV latency, the frequency of infected cells has been estimated to be 800/10^6^ cells among immune suppressed patients^15^, and EBV is not latent in T cells. The changes in multiple cell populations imply that there are bystander effects and suggests that either soluble mediators may be produced in response to the prior infection or that T/B cell interactions are affected. Several mechanisms may be postulated: SoRelle et al described a model in which EBV-infected B cells continuously drives recurrent B cell entry, progression through, and egress from the Germinal Center (GC) reaction creating a “perpetual GC”. This recurrent cell activation may have effects on both T and B cells and has been associated with features of B cells in autoimmune disorders^16^. BCRF1 of EBV encodes a viral IL10 homologue (vIL10), EBV produces human IL-10 and also encodes a decoy vCSF1R, which binds CSF1 and thereby limits mobilization of hemopoietic stem cells^17, 18^.The EBV-encoded IL-10 has weaker functionality than cellular IL-10 t (cIL-10), but it may be additive or synergistic with cellular IL-10 that is induced by teplizumab and thereby inhibit cellular immune responses ^19^. EBV downregulates Class I MHC and interferes with presentation of viral peptides on Class I and Class II MHC *via* BDLF3-induced ubiquitination and by BNLF21 by preventing Class I MHC peptide loading by inhibiting the transporter associated with peptide loading (TAP)^17^. gP42 can be released in a soluble form, which inhibits interaction between Class II MHC and the T cell receptor ^20, 21^.The EBV protein, latent membrane protein 2A (LMP2A), co-opts tyrosine kinases used by the T cell receptor ^22^. Stable expression of LMP2A in Jurkat T cells down-regulated T cell receptor levels and attenuated T cell receptor signaling. EBV peptides can bind to HLA-E which is a ligand for NKG2A ^23^. In previous studies we showed that NKG2A that was induced with Teplizumab treatment, served as a ligand for activated CD8^+^ T cells with regulatory function^24^. Finally, Hong et al identified more than 1700 regions in the human genome where EBNA2 altered chromatin looping interactions and Harley et al found that EBNA2-anchored genetic associations exist in multiple autoimmune diseases ^25, 26^.

In addition to the effects of prior EBV infection on CD8^+^ T cells we identified differences in CD4^+^ T cells, B cells, and Tregs. Curiously, the timing of effects on cell subsets differed. For some CD8^+^ T cells there were significant differences before treatment whereas for the others the differences were detected even 18 months after teplizumab. For CD4^+^ cells the differences at the baseline best discriminated EBV seropositive and EBV seronegative individuals. The changes in B cells were more proximal to the timing of treatment.

The effects of latent EBV may depend on disease specific autoimmune mechanisms. Several autoimmune diseases including MS, Sjogren’s syndrome, rheumatoid arthritis, and SLE have been related to EBV infection. Recently, Lanz et al found clonally expanded B cells bind EBNA1 and GlialCAM in patients with MS suggesting that there is cross reactivity between viral proteins and autoimmune targets^6^ Both a reduced content and dysfunction of Tregs are closely related to the occurrence and development of SLE^27^. Our studies are unique since EBV is not thought to have a causative role in T1D and because the study participants were relatively young, there was a large proportion of EBV seronegative patients. Furthermore, we were able to evaluate the functional effects on immune cells after teplizumab.

There are limitations to our studies. In both trials the frequency of EBV seropositive and seronegative participants was not equally balanced and there were also more CMV seronegative participants. This may reflect the inclusion of pediatric patients who were more frequently seronegative for both viruses. Because the frequency of EBV seropositive increased with age, we were unable to distinguish the effects of age in addition to just EBV serostatus. Further studies with larger numbers of older EBV seronegative individuals would help to clarify this potential confounder. Among the 17 EBV seropositive individuals, 12 were also CMV seropositive and the combined effects of the latent viruses may have affected the findings with EBV. In addition, as noted above, we cannot exclude that there may have been reactivation of EBV that was not captured by measuring of viral loads in the study protocols or given the length of both of these studies, additional infections with EBV or CMV may have occurred in baseline seronegative participants during the study course.

In summary, we have identified how prior infection with EBV has broad effects on the immune repertoire and responses to a biologic treatment. Our findings may have important implications for understanding the development of autoimmune diseases. These findings may also help to develop personalized approaches to immune therapy that consider the biologic activity of the agent in the setting of the host.

## Materials and Methods

### Clinical studies

Samples and clinical data were collected from two randomized clinical trials of teplizumab that have been described previously ^8, 28^. In the TN10 trial, relatives without the diagnosis of clinical T1D were identified as high-risk for the diagnosis of clinical disease on the basis of positive autoantibodies and dysglycemia (i.e. Stage 2 T1D). Their median age was 14.0 years (range 8.5-49.1 years). They were randomized to treatment with a single 14-day course of teplizumab or placebo. The participants were followed at approximately 6 months intervals for the diagnosis of clinical, Stage 3 T1D, the primary endpoint, using oral glucose tolerance tests (OGTT). Samples for mechanistic studies were collected before treatment and at approximately 3, 6, and 18 months after treatment.

In a second study (AbATE, ITN027AI), patients diagnosed with T1D (median age 12.1 years, range 8.2 to 29.6 years) were randomized to treatment with a 14 day course of teplizumab or to an observation group. A second course of teplizumab was given at 12 months. Samples were collected at similar intervals. Metabolic responses to a mixed meal tolerance test (MMTT, C-peptide secretion) were measured at 6 months intervals and the stimulated C-peptide levels between the treatment arms and viral serology categories, corrected for the baseline levels, was assessed.

In both studies, participants underwent screening for EBV (anti-EBVIgG, anti-EBNA, and/or anti-EBV IgM) and CMV (anti-CMV IgG, IgM) at the time of enrollment. EBV and CMV viral copies were also measured during the studies (Table 1). All patients provided written informed consent or assent. The studies were approved by the Institutional Review Boards at the study sites.

The Institutional Review Boards at the following locations gave ethical approval: For the TN10 protocol: Children’s Mercy Hospital, Overland Park, KS USA Vanderbilt University, Nashville, TN USA, University of Florida, Gainesville, FL USA, Yale University, New Haven, CT USA, University of Miami, Miami, FL USA mBarbara Davis Center, University of Colorado, Anshutz, CO, USA, University of Minnesota, Minneapolis, MS USA, University of California-San Francisco, San Francisco, CA USA, Hospital for Sick Children, Toronto Canada, University of Iowa, Iowa City, IA USA, University of South Florida, Tampa, FL USA, Forschergruppe Diabetes, Klinikum rechts der Isar der Technischen Universitat Munchen, Munich, Germany. For the AbATE protocol: Yale University, New Haven, CT USA, University of California San Francisco, San Francisco, CA USA, University of Colorado, Anshutz, CO USA, Benaroya Research Center, Seattle, WA, University of Washington, Seattle, WA. All study participants gave written consent or assent for use of their samples for mechanistic studies.

### Metabolic assessments and flow cytometry

In the TN10 trial, the progression from Stage 2 to Stage 3 T1D was assessed using confirmed responses to an oral glucose tolerance test. In AbATE, the stimulated C-peptide levels were measured during a 4 hr mixed meal tolerance test (MMTT). The AUC was calculated and for analysis, transformed (ln (AUC+1)/240). OGTT C-peptide and glucose values were tested by Northwest Lipids Research Laboratories using the TOSOH C-peptide immunoassay and Roche glucose assay. EBV serologies were measured at the University of Colorado (TN10) or at ViraCor (AbATE). PBMCs were processed and stored at the NIDDK or ITN repository. Cryopreserved vials of PBMC were sent to ITN Core laboratory at Benaroya Research Institute for analysis by flow cytometry with antibody panels as described previously described previously ^11, 28^.

### Single cell RNA sequencing processing and analysis

CellRanger was utilized to process the raw sequence data. The gene-cell barcode matrices were used for further analysis with the R package Seurat (Seurat development version 4.0.2) with additional utilization of the packages dittoSeq, harmony, scCustomize, SCpubr, monocle3, pheatmap, EnhancedVolcano. Demultiplexing was done using HTODemux with automatic thresholding. Cells were filtered if they were classified as doublets or negative for hashtag antibody based on the demultiplexing results, or if they had fewer than 200 features or greater than 5% mitochondrial RNA was detected.

After removing likely multiplets and low-quality cells, the gene expression levels for each cell were normalized with the NormalizeData function in Seurat followed by the integration of the single cell data. The integrated data was scaled and the principal analysis was performed. Clusters were identified using FindNeighbors and FindClusters Seurat’s functions. Batch correction was applied using RunHarmony function. Cell cluster identities were manually defined with the cluster-specific marker genes or known marker genes. The cell clusters were visualized using Uniform Manifold Approximation and Projection (UMAP) plots and complete using plot3D. CD8^+^, NK, CD4^+^, B cells, monocytes and dendritic cells, were re-clustered separately at a resolution between 0.1-0.2 to obtain biologically meaningful clusters each respectively. Subsets functions were used to compare the different types of cells included in the study. A MAST package to run the DE testing implemented in the FindAllMarkers function in Seurat was used to identify up regulated and down regulated genes associated with each individual subset and used for later analysis. IPA software was used to analyze pathway expression. To investigate the kinetics of gene expression during T and B cell differentiation, we performed single-cell trajectory analysis using the Monocle 3 package. The scRNA-seq profiles of CD8^+^, CD4^+^ and B cells were used to reconstruct the single-cell trajectories for the different states. The group-specific marker genes were selected using the “graph_test” function. We pseudo-temporally ordered the cells using the“reduceDimension” and “orderCells” functions. The significance of upregulated expression in the cells was tested by Moran’s I test available in graph_test function.

### B cell stimulation and Western blots

B cells were purified with magnetic separation using CD20 microbeads (Miltenyi Biotec). B cells were plated at 100.000 cells/well in a 96-well plate in RPMI/10% FBS and 2.5 μg/ml polyclonal F(ab′)2 anti-human IgM (Jackson Immunoresearch).

For Western blot, pre-stimulated purified B-cells were lysed, and proteins extracted using 1X Cell Lysis Buffer (Cell Signaling #9803) supplemented with protease and phosphatase inhibitor cocktail (Thermo Fisher, Cat#78440). A total amount of 10 ug total protein per patient sample was separated by SDS-PAGE and then transferred to a nitrocellulose membrane (Bio-Rad). Signals were detected using SuperSignal West Pico PLUS (Thermo Fisher, Cat#34579) chemiluminescent substrate. The primary antibodies and dilutions used for immunoblotting were as follows: pERK1/2 (Cat#4370, 1:1000), ERK1/2 (Cat#4695; 1:1000), ERK1/2 (Cat#4695; 1:1000) and ACTIN (Cat#4970; 1:1000). Secondary antibody anti-rabbit HRP (Cat#7074; 1:2000). Primary and secondary antibodies were from Cell Signaling.

### Assays

Glucose and C-peptides were measured at the Pacific Northwest Lipid Laboratory, Seattle WA, the latter with a TOSOH assay described previously^11, 28^.

### Statistical analyses

The effects of drug treatment were compared in patients based on their EBV or CMV serostatus at the time of study enrollment. The rates of conversion from Stage 2 to Stage 3 T1D in the TN10 trial were compared by Log-Rank test with Sidak’s adjustment for multiple comparisons. For AbATE, repeated measures analysis of variance was performed with a mixed model using SAS 9.4 (Cary, North Carolina) with adjustment for baseline C-peptide levels but without adjustment for the baseline for immunologic measures. The frequencies of cell subsets were analyzed with a mixed model SAS 9.4 without correction for the baseline values. Multiple t-tests were used to compare cell subsets by flow cytometry was performed with GraphPad Prism 9 software with FDR correction. The data were log transformed for analysis.

## Data Availability

All data produced in the present study are available upon reasonable request to the authors

## Acknowledgements

KH is a co-inventor on a patent for the use of teplizumab for delay of Type 1 diabetes. Supported by grants DK057846 and AI66387 to KH. Research reported in this publication was supported by the National Institute of Allergy And Infectious Diseases of the National Institutes of Health under Award Number UM1AI109565. The content is solely the responsibility of the authors and does not necessarily represent the official views of the National Institutes of Health

---

**Supplemental Figure 1.**
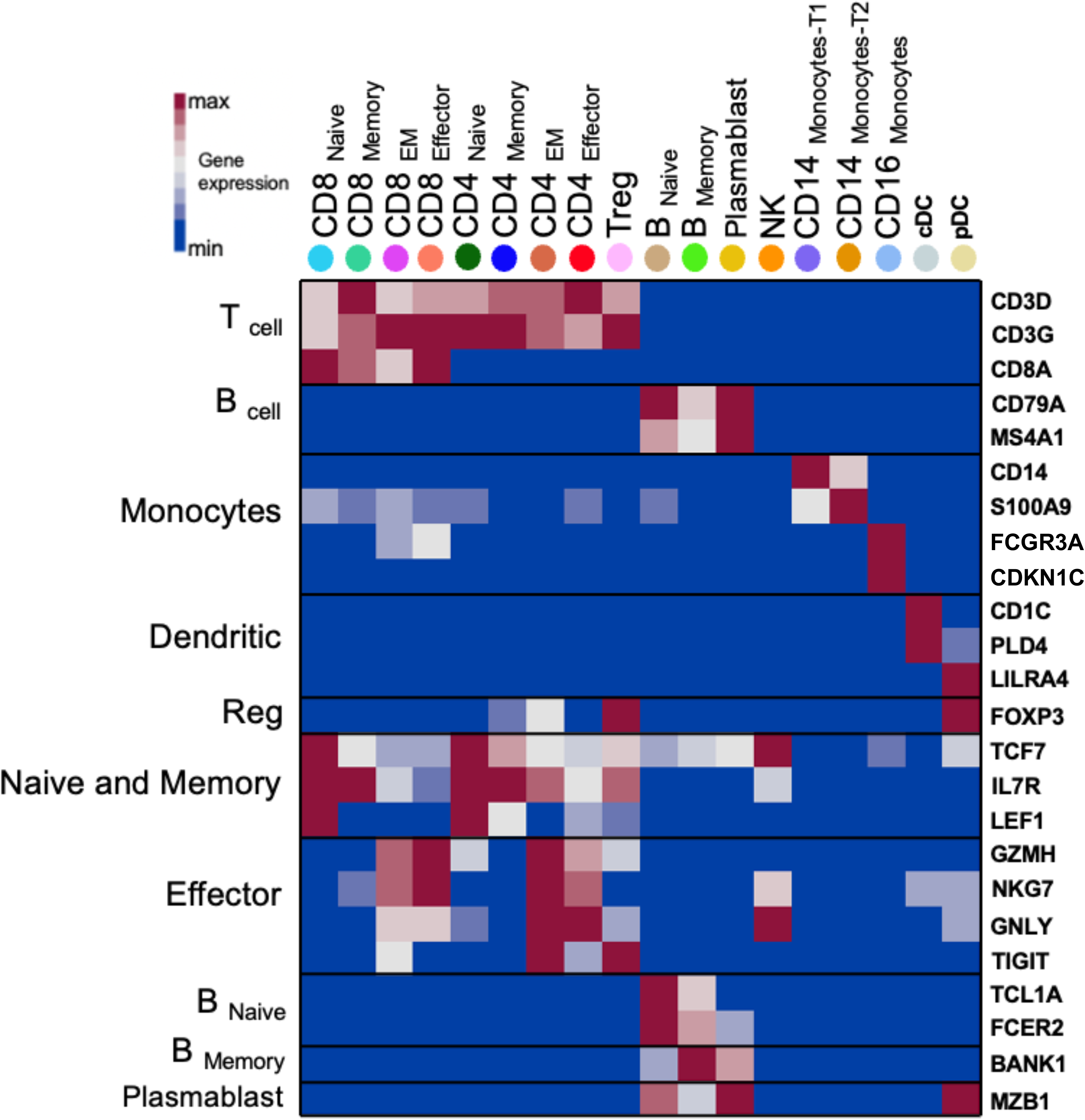
Heatmap showing gene expression of selected markers used for cluster annotation. cDC: classical dendritic; pDC: plasmacytoid dendritic.

**Supplemental Figure 2.**
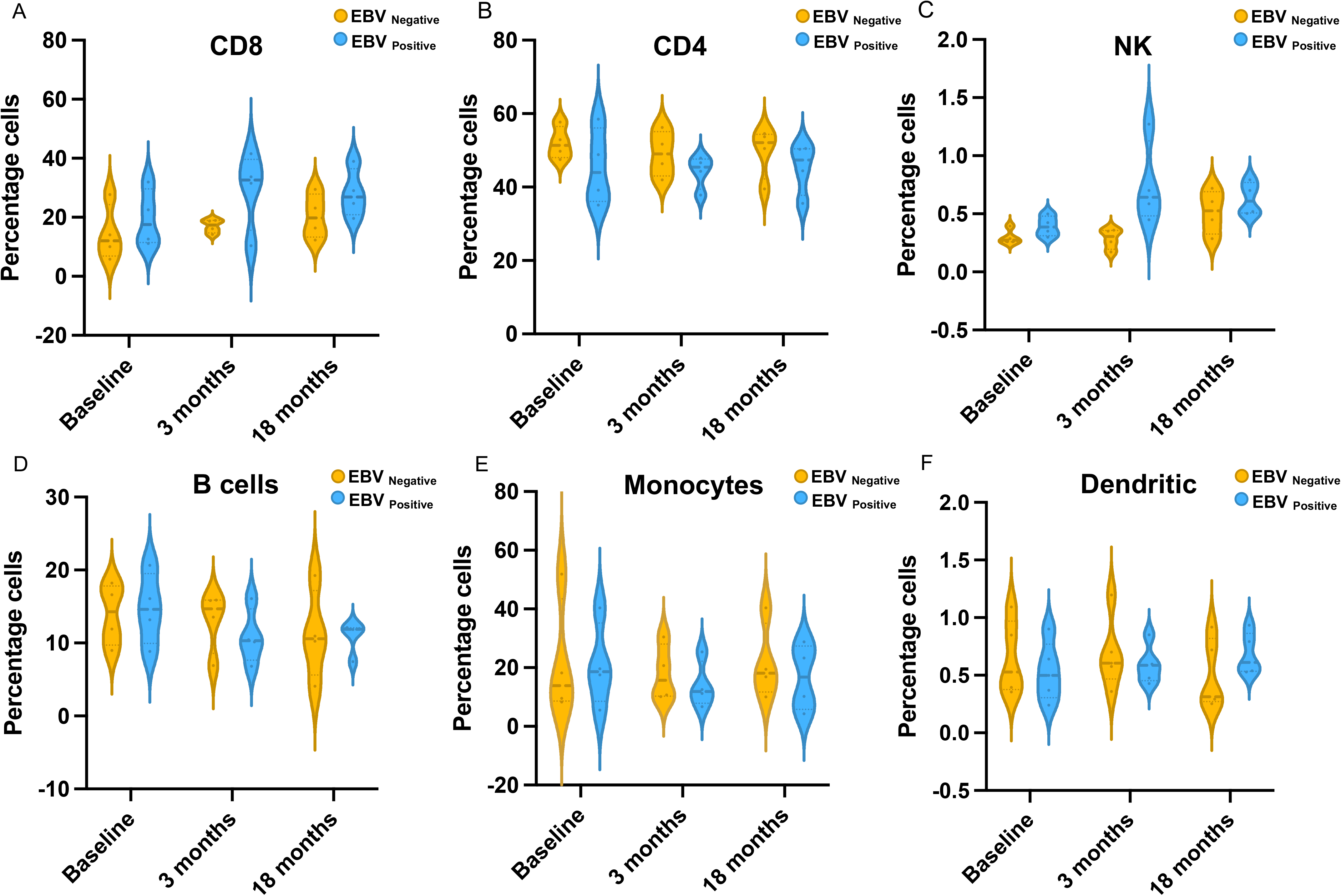
(A-F) Violin plots showing the percentages of CD8^+^, CD4^+^T cells, NK, B cells, monocytes and dendritic cells at the study visits obtain by scRNA-seq. Color denote EBV serostatus.(n=4 EBVseropositive, n=4 EBVseronegative)

**Supplemental Figure 3:**
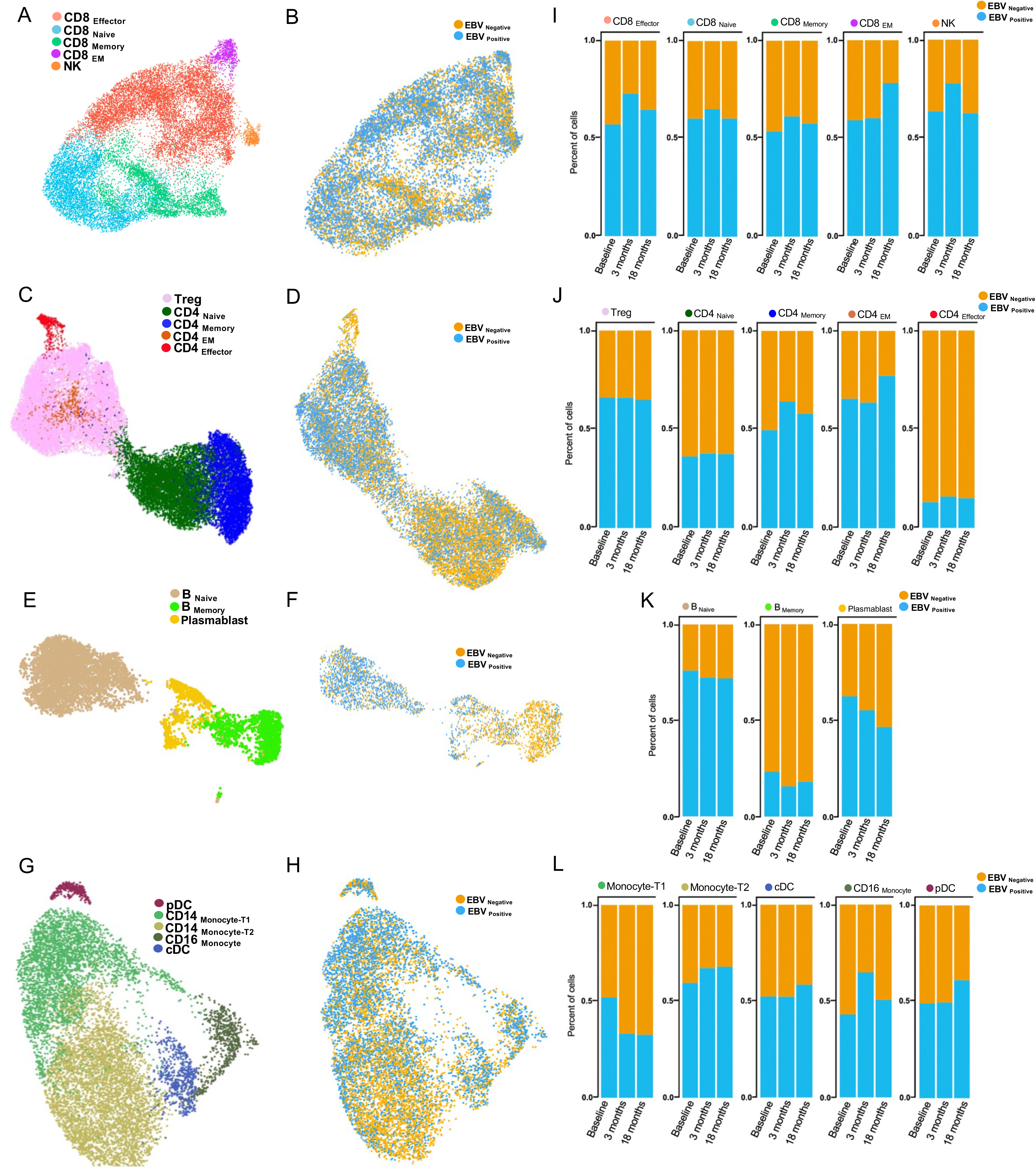
Subclusters distribution and differences between EBV positive and negative participants. (A,C,E,G) UMAPs of CD8^+^, CD4^+^ T cells, B cells, and monocytes and dendritic cells respectively. Points represent individual cells and color denote cluster classification as labeled. (B,D,F,H) UMAPs visualization of CD8^+^, CD4^+^ T cells, B cells, and monocytes and dendritic cells respectively by EBV serostatus. Points represent individual cells and color denote EBV classification as labeled. (I, J, K, L) Bar plots illustrating the percentage of cells in the CD8^+^, CD4^+^ T cells, B cells, and monocytes and dendritic cells respectively in the teplizumab treated patients (n=8). The percentage (of the total number of cells for the EBV seronegative (orange= (n=4)) and EBV seropositive (blue, n=4)) for each participant was calculated. The bars show the average proportion of the cells based on serostatus. pDC: plasmacytoid dendritic; cDC: classical dendritic

**Supplemental Figure 4.**
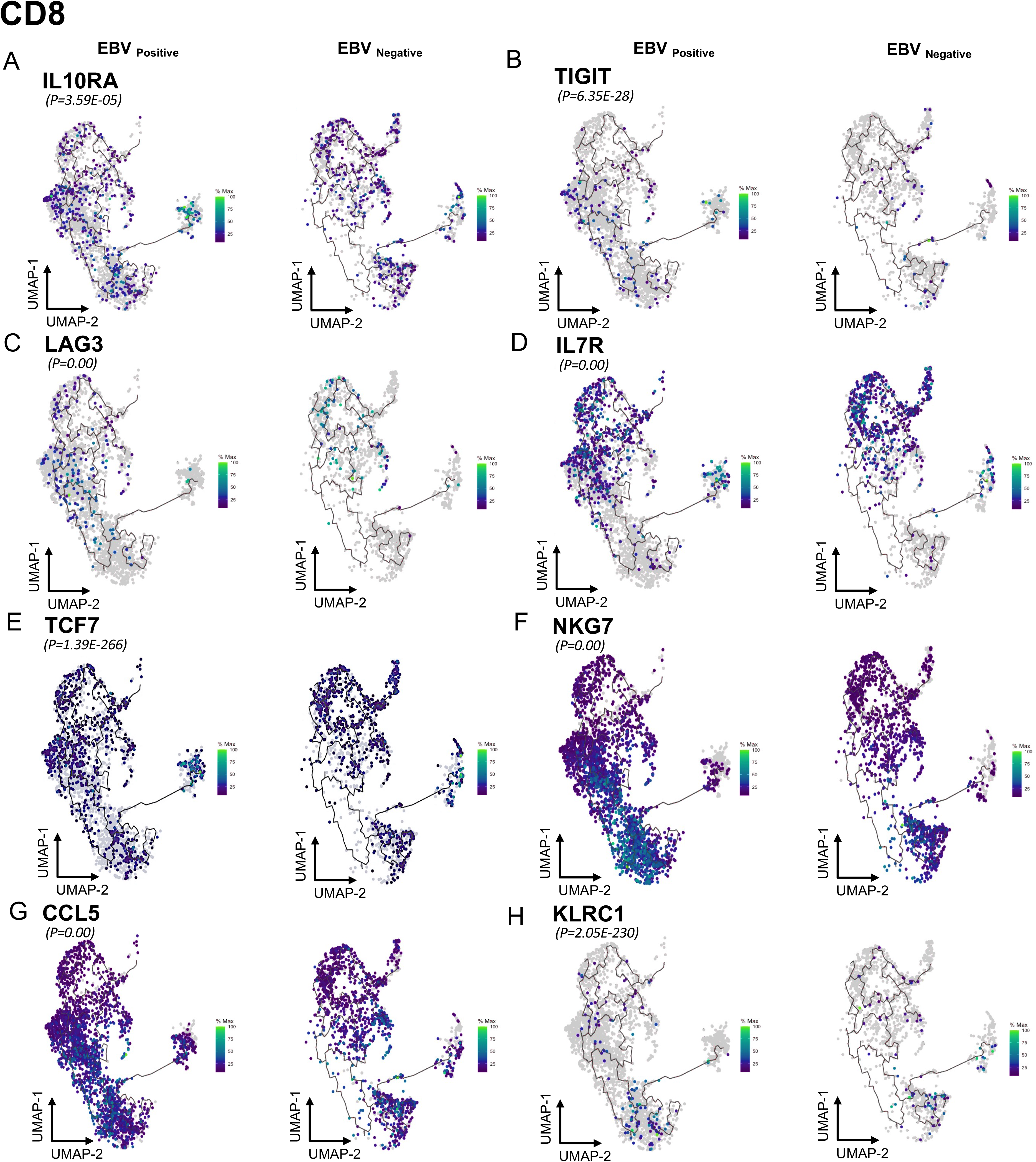
(A-P) UMAPs showing the expression of TCF7^+^, LEF1^+^, IL7R^+^, NKG7^+^, GZMK^+^, CCL5^+^, CTLA4^+^, NR4A1^+^ in different stages of differentiation of CD4^+^ T cells in the EBV seropositive vs EBV seronegative under teplizumab at 18 months. P value indicates significative change across the pseudotime (n=4 EBV seropositive, n=4 EBV seronegative).

**Supplemental Figure 5.**
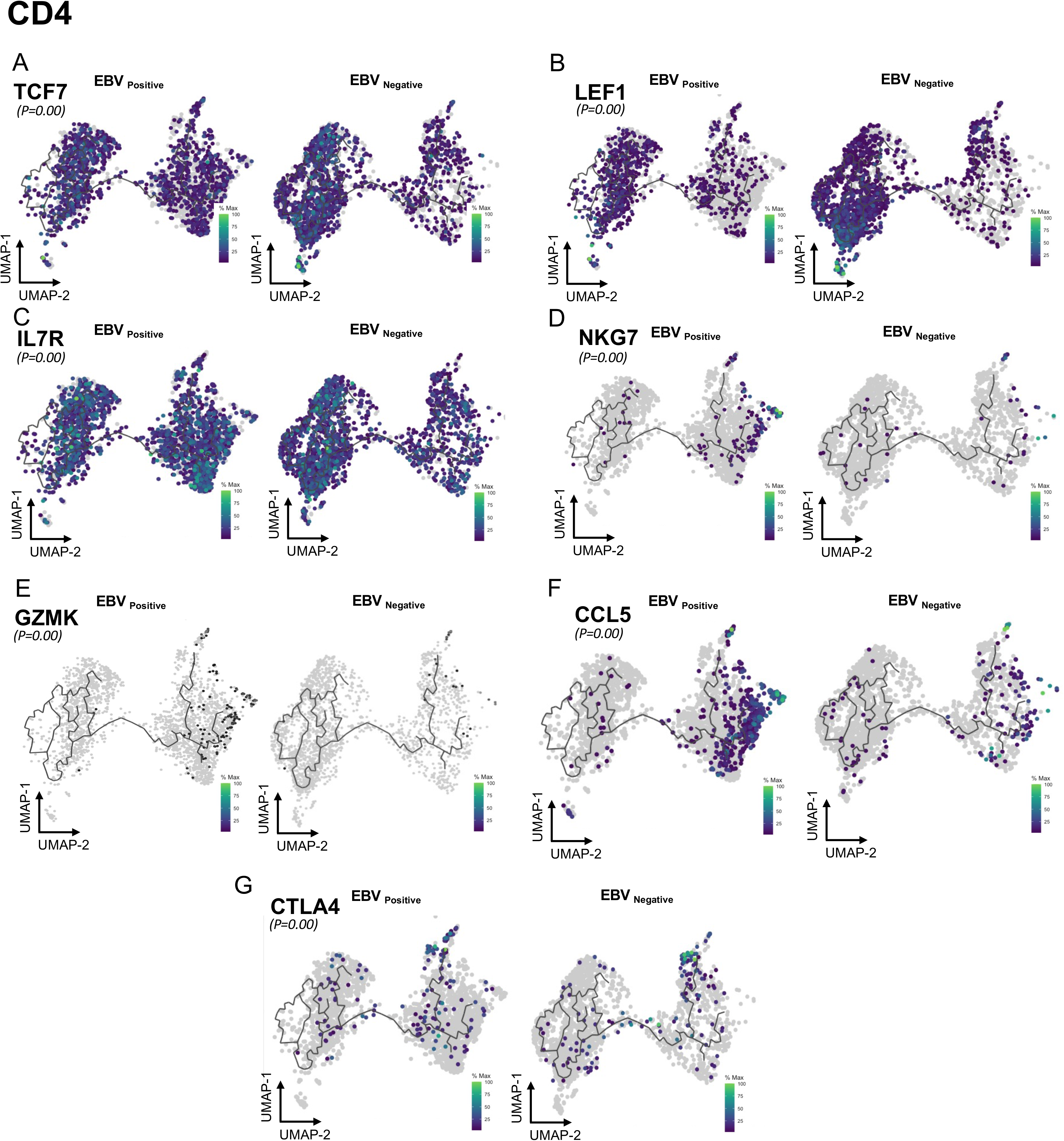
(A-O) UMAPs showing the expression of IL10RA^+^, TIGIT^+^, LAG3^+^, IL7R^+^, TCF7^+^, NKG7^+^, CCL5^+^, KLRC1^+^ in different stages of differentiation of CD8^+^ T cells in the EBV Positive vs EBV negative under teplizumab at 18 months. P value indicates significative change across the pseudotime. (n=4 EBV seropositive, n=4 EBV seronegative).

**Supplemental Figure 6.**
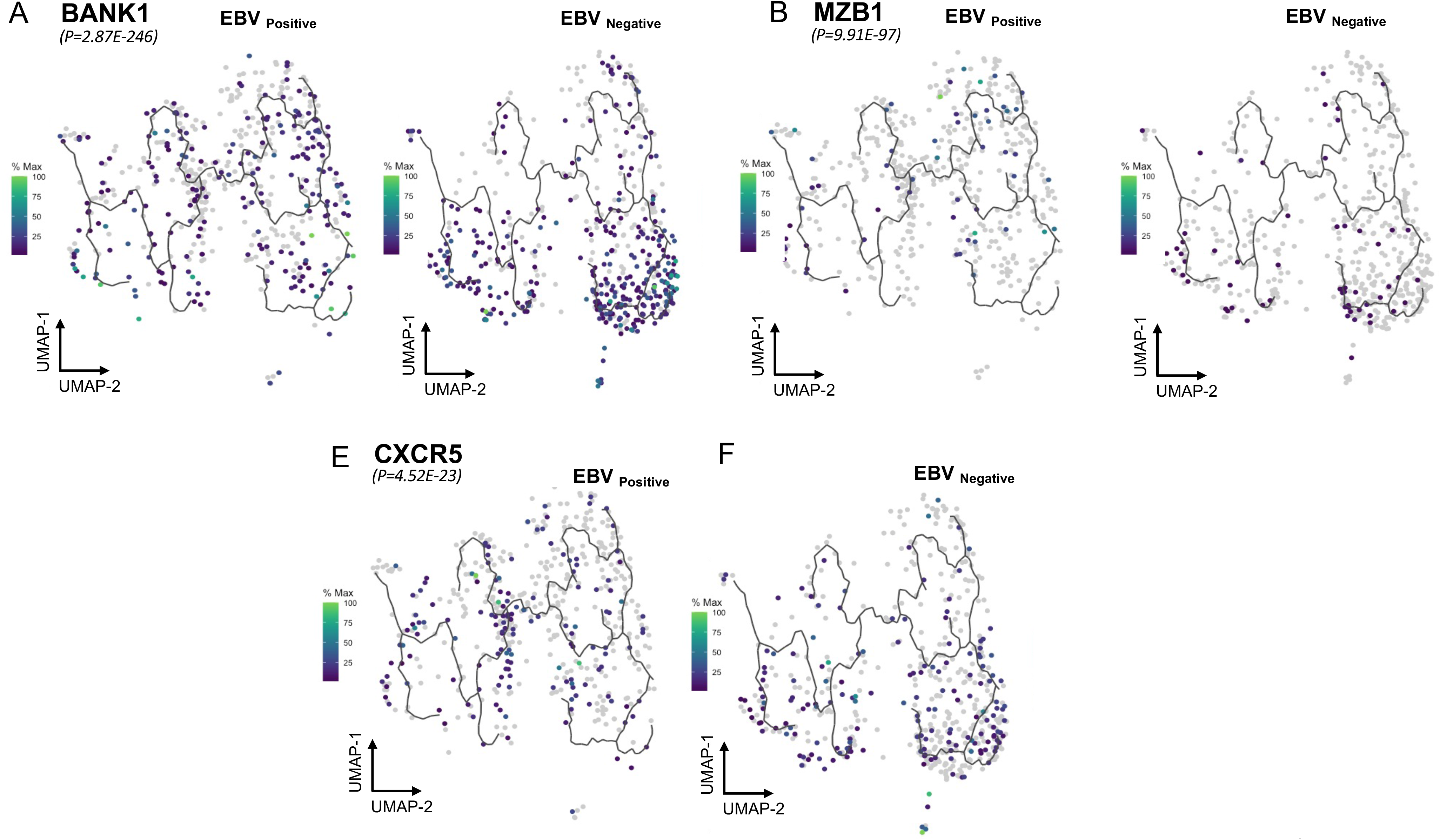
(A-F) UMAPs showing the expression of BANK1^+^, MZB1^+^, CXCR5^+^ in different stages of differentiation of B cells in the EBV seropositive vs seronegative under teplizumab at 18 months. P value indicates significative change across the pseudotime. (n=4 EBV seropositive, n=4 EBV seronegative).

